# Spinal fluid antibodies against multiple sclerosis candidate bacteria in demyelinating disease

**DOI:** 10.1101/2021.02.05.21250635

**Authors:** Emily Eckman, Jon D. Laman, Kael F. Fischer, Bert Lopansri, Tom B. Martins, Harry R. Hill, John D. Kriesel

## Abstract

A panel of 10 IgG enzyme-linked immunosorbent assays (ELISAs) were developed for the detection of anti-microbial immune responses in the cerebrospinal fluid (CSF) of patients with demyelinating diseases (DD). Selection of the anti-microbial ELISA assays was guided by previous RNA sequencing studies that established a multiple sclerosis (MS) microbial candidate list. Microbial antigens included on the ELISA panel were derived from *Akkermansia muciniphila, Atopobium vaginae, Bacteroides fragilis, Lactobacillus paracasei, Odoribacter splanchnicus, Pseudomonas aeruginosa, Cutibacterium (Propionibacterium) acnes, Fusobacterium necrophorum, Porphyromonas gingivalis*, and *Streptococcus mutans*. Spinal fluid responses from patients with demyelinating diseases (DD, N=14) were compared to those with other neurological diseases (OND, N=8), and non-MS (Control, N=13) control patients. Commercial positive and negative control CSF specimens were run with each assay. ELISA Index values were derived for each specimen against each of the 10 bacterial antigen preparations. Intrathecal production of anti-microbial antibodies was assessed by comparing CSF and comparably diluted serum. CSF reactivity was significantly higher in the DD group compared to the controls against Akkermansia, Atopobium, Bacteroides, Lactobacillus, Odoribacter, and Fusobacterium. Four of the 11 tested DD group subjects had elevated antibody indexes against at least one of the 10 bacterial antigens, suggesting intrathecal production of anti-bacterial antibodies. This CSF serological study supports the hypothesis that several of the previously identified MS candidate microbes may contribute to demyelination in some patients.

## INTRODUCTION

A number of infections of the central nervous system (CNS) can lead to demyelination, including distemper (dogs), measles (humans), JC virus (human), influenza (humans), and now possibly SARS-CoV-2.^1, 2^ The underlying risk factors and mechanisms for microbe-driven demyelination are poorly understood, since it occurs unpredictably and at variable rates.

Microbes, particularly herpes viruses such as EBV and HHV6, have long been suspected as causative agents of the prototypic demyelinating disease multiple sclerosis (MS), based on the epidemiology of the disease including geographic patterns, isolated outbreaks, and migration studies.^3, 4^ However, this field is contentious with much conflicting evidence. Establishment of causal relationships is complicated by the prolonged time interval (years to decades) between initial exposure, likely in puberty, and the development of MS, most often in young adulthood. Researchers in the 1970s and 1980s made extensive efforts to find and isolate possible viral pathogens from fresh autopsy brain tissue.^5^ These efforts included inoculation of diseased human brain tissue into cell cultures, live animals, and even eggs, without any evidence of replicating viruses. However, bacterial and fungal cultures were not performed, and ex vivo viral culture techniques included the use of antibiotics in the culture medium. While these efforts to find viruses responsible for MS were certainly ambitious and laudable, this strategy likely would not have detected most bacteria, fungi, or protists within the affected brain tissue.

We recently utilized next-generation unbiased RNA sequencing of demyelinated human brain samples from living subjects as a method for defining possible microbial contributors to MS and related demyelinating diseases (e.g. acute disseminated encephalomyelitis, ADEM; neuromyelitis optica, NMO).^6^ RNA sequencing of fixed, paraffinized brain biopsy specimens from these research subjects was performed in comparison to control epilepsy brain samples, including blank samples to rigorously control for environmental and reagent contamination. This work showed significantly more mapped microbial sequences in the demyelinated brain samples than in the controls. Stringent computational analysis of the sequencing data resulted in a list of 29 differentially expressed MS microbial candidates, mostly anaerobic bacteria which are often difficult to culture. Some of these bacterial species are derived from the physiological microbiota of the human gut and vaginal epithelium.

Collectively, these findings suggest that both exogenous bacterial pathogens and commensal species could directly or indirectly contribute to the development and progression of demyelinating disease. This study was designed to determine if some MS candidate microbes, discovered by sequencing of brain tissue, are also eliciting an immune response in the CSF of patients with demyelinating diseases, including MS. Ten organisms were selected for further study. Bacterial lysates were used to develop a panel of 10 novel dedicated ELISA assays. The ELISA panel was applied to CSF from patients with demyelinating disease (DD), other neurologic disease (OND), and controls. The data demonstrates significantly higher CSF reactivity among DD subjects against several of the panel components compared to controls.

## MATERIALS AND METHODS

### Subjects, CSF and Serum Collections

The study was approved by the University of Utah Health Sciences Institutional Review Board (IRB). Each subject either signed an IRB-approved informed consent or had a legally authorized representative sign for them. CSF was obtained from 14 patients with MS or other demyelinating diseases (DD) during the course of their regular clinical care. CSF was also collected from 8 patients with other neurologic diseases (OND) and 13 controls. Most of the OND subjects were enrolled in the study for suspicion of demyelinating disease, but the final neurologic diagnosis was something else (e.g. vasculitis). The controls were patients undergoing spinal fluid shunt placements or exchanges who did not have active infection at the time of collection. These control subjects did not have clinical tests performed on their CSF samples, but were expected to have normal CSF based on their stable clinical condition. Paired serum samples taken around the time of CSF collections were obtained from 11 DD and 8 OND subjects. Serum was not collected on the control cohort. Oligoclonal band OCB testing and albumin index determinations were performed (ARUP Laboratories) as part of regular clinical care in some of the DD and OND subjects. CSF and serum IgG concentrations were determined by nephelometry, also at ARUP laboratories.

### Criteria for microbial selection and inclusion

Selection of microbial candidates was guided by the MS microbial candidate list. ^6^ MS candidate microorganisms were selected for optimization if they were: 1) readily available from ATCC or clinical sources, 2) culture conditions were not too demanding or dangerous, and 3) they were widely distributed among the contributing microbial families or were of particular interest. Characteristics of and justification for the microbes selected for the CSF serology panel are displayed in Table 1. The organisms were grown in the appropriate conditions, washed, diluted to optical density (OD) of approximately 0.5 or 1.0 (∼10,000 cells/ml), and sonicated. Microbes selected for the CSF serology panel include *Akkermansia muciniphila* (ATCC BAA-835), *Atopobium vaginae* (ATCC BAA-55), *Bacteroides fragilis* (ATCC 29771), *Cutibacterium (Propionibacterium) acnes* (ATCC 6919), *Lactobacillus paracasei* (ATCC 27092), *Odoribacter splanchnicus* (ATCC 29572), *Porphyromonas gingivalis* (ATCC 33277), *Pseudomonas aeruginosa* (ATCC 10145), *Streptococcus mutans* (ATCC 25175) and *Fusobacterium necrophorum* (ATCC 25286).

**Table 1.** Characteristics of the 10 bacteria selected for the CSF ELISA panel.

| Organism | Ecology | Source<sup>1</sup> | MS:control Ratio<sup>2</sup> | Justification for Inclusion |
| --- | --- | --- | --- | --- |
| *Akkermansia muciniphila* | gut anaerobe | ATCC BAA-835 | 81 | RNA-seq candidate, MS association |
| *Atopobium vaginae* | vaginal anaerobe | ATCC BAA-55 | 8 | RNA-seq candidate, normal vaginal flora |
| *Bacteroides fragilis* | gut anaerobe | ATCC 29771 | 19 | RNA-seq candidate |
| *Lactobacillus paracasei* | vaginal aerobe | ATCC 27092 | 7 | RNA-seq candidate |
| *Odoribacter splanchnicus* | oral anaerobe | ATCC 29572 | 74 | RNA-seq candidate |
| *Pseudomonas aeruginosa* | waterborne aerobe | ATCC 10145 | Infinite<sup>3</sup> | RNA-seq candidate, host of bacteriophage Luz24likevirus |
| *Cutibacterium (Propionibacterium) acnes* | skin anaerobe | ATCC 6919 | 4 | RNA-seq candidate |
| *Fusobacterium necrophorum* | oral anaerobe | clinical isolate | 13 | RNA-seq candidate |
| *Porphyromonas gingivalis* | oral anaerobe | ATCC 33277 | 4 | RNA-seq candidate, Alzheimer's association |
| *Streptococcus mutans* | oral aerobe | ATCC 25175 | 13 | RNA-seq candidate |

<sup>1</sup> ATCC catalog numbers provided (https://www.atcc.org)
<sup>2</sup> (Sum mapped read pairs in the MS group) divided by (Sum mapped read pairs in the Control group) (see Table S3)
<sup>3</sup> Refers to sequencing results for the Luz24likevirus: 8,338 mapped reads in the MS group and zero mapped reads in both the control and blank specimens

### Bacterial antigen preparation

Bacterial strains were obtained from ATCC as freeze-dried, lyophilized cultures. Cultures were rehydrated using the appropriate medium and incubation conditions specified on the product sheets. Organisms were allowed to come out of the lyophilized state, and once within the exponential phase of growth, stock cultures were stored at −80 degrees C. in 10% glycerol. Broth and plate cultures intended for antigenic harvesting were harvested during the exponential phase or shortly thereafter. Prior to harvesting antigen, cells from broth cultures were washed with PBS at least twice, and serial dilutions were conducted from both plate and broth cultures in order to obtain an OD of 1.0 using ELISA coating buffer (Supplier), equivalent to 3.9 × 10^8^ cells per milliliter and 1.93 × 10^7^ cells per well in a flat-bottom 96-well plate (Corning Life Sciences Product 9018). Plates were read on a Gen5 2.08 ELISA plate reader at 600 nm. Cells were sonicated for 10 seconds with cycle time continuous, a duty cycle of 40%, and at position 4.5 with a Branson Sonifier 250. The sonicated bacterial antigens were aliquoted and stored at −80 degrees C. for future use.

### ELISA Procedure

Flat-bottom 96-well ELISA plates were coated with 50µl/well of harvested antigen (sonicated bacteria), covered, and rocked overnight at 4 degrees C. The plates were washed with phosphate buffered saline with 0.1% Tween 20 (PBST), blocked with Starting Blocker for 1 h at room temperature, and washed again before applying the appropriate primary antibody for 2 h. Secondary antibodies were applied for 1 hr. The plates were developed with TMB for five min and stopped with 2N HCL. Absorbance was read on a Gen5 2.08 ELISA plate reader at 450nm.

A series of steps were taken to optimize the 10 separate ELISA assays. Four commercial blocking agents were assessed for blocking efficiency including 5X ELISA/ELISPOT Diluent (Invitrogen), Blocker BLOTTO in tris-buffered saline (ThermoScientific), Blocker Casein (ThermoScientific), and Starting Blocker (ThermoScientific). Starting Blocker proved to be optimal for most of the assays, and was adopted for use with all 10 bacterial antigens. Three primary anti-bacterial antibodies were assessed as positive controls for assay performance validation: mouse monoclonal IgG3 anti-peptidoglycan (MAB995, Sigma-Aldrich), mouse monoclonal IgG2b anti-LPS (ab35654, Abcam), and rabbit polyclonal anti-pseudomonas (PA1-73116, Thermo Fisher Scientific). These positive control primary Ab’s were titrated to yield strong and weak (just above the background) positive signals. The polyclonal anti-Pseudomonas Ab PA1-73116 proved to be the best primary positive control antibody against all the bacterial antigens tested. This positive control Ab was used at dilutions between 1:200 and 1:6400 as strong (OD ∼1.0) and between 1:400 and 1:102,400 as weak (OD > background) positive controls for each of the 10 assays. Secondary antibodies used in the study included anti-mouse IgG (Vector PI-2000, 1:3000), anti-rabbit IgG (Vector PI-1000, 1:3000), and anti-human (Jackson ImmunoResearch, 1:10,000 or 1:20,000). The antibodies and dilutions used for the study are detailed in Table S1.

Commercial human CSF (Randox Laboratories Ltd, UK; ([IgG] 10 mg/dl) was used as a positive control. IgG-depleted commercial human CSF (Randox Laboratories Ltd, Crumlin, UK) was utilized as a negative (calibration) control. The “cut-off calibrator” method was chosen for each of the assays.^7 8, 9^ This involved making calibrator (negative) control CSF samples depleted of virtually all IgG. Effective IgG depletion was performed with two elutions of commercial CSF (Randox®) through the HiTrap Protein G HP kit (GE Healthcare, Product 29-0485-81). IgG depletion was verified with Human Total IgG Platinum ELISA kit (Affymetrix eBioscience). The final depleted IgG concentration was 5.25 ng/ml. Other negative controls run with each assay included PBS only (blank) and (no-primary) secondary Ab only wells.

### Analysis of ELISA Data

CSF and diluted serum samples were run in duplicates. An average OD was taken for each experimental and control sample and divided by the cutoff calibrator to obtain an ELISA Index (EI) value for each of the samples. Matched serum samples were collected from as many of the DD and OND subjects as possible. CSF and serum IgG levels were obtained from clinical data, and/or from nephelometry determinations, all performed at ARUP Laboratories (Salt Lake City, Utah, USA). The serum samples were diluted in PBS to match each sample’s CSF IgG concentration. This allowed the direct assessment of intrathecal IgG synthesis using the EI_CSF_:EI_serum_ ratio or Antibody Index (AI). An AI greater than one shows excess Ab in the CSF compared to serum, suggesting that intrathecal Ab synthesis has taken place. ^10, 11^

### Statistical Analysis

The ages of the subjects in the DD, OND, and control groups were compared by one-way ANOVA, and sex distributions of the subjects were compared using the 2×3 Fishers Exact test.^12^. EI values among the groups were screened with an unweighted one-way ANOVA.^12, 13^ Differences in EI values between the groups were compared by using 2-tailed Mann-Whitney nonparametric testing.^14^ EI’s and albumin index values were compared by linear regression, performed within the Prism 9.0 computer application.^13^

## RESULTS

### Study population

The characteristics of the study population are shown in Table 2. Of the 14 patients that were diagnosed with demyelinating disease (DD), 2 had relapsing-remitting MS, 1 had a clinically isolated syndrome, 3 had tumefactive MS, 3 were classified as untyped or neurodegenerative MS, 4 had ADEM, and 1 had anti-MOG syndrome. OND subjects had CNS Lyme Disease, vasculitis, rhombencephalitis, neurosyphilis, migraine, spina bifida, atypical stroke, CSF shunt infection with *C. acnes*, or pudendal neuralgia. Median ages of the subjects were 43 (DD), 42 (OND), and 62 (Control), not significantly different (p = 0.23). The sex distributions of the groups were also not significantly different (p = 0.24).

**Table 2.** Characteristics of the Study Subjects.

| Group-Specimen | Age | Sex | Neurologic Diagnosis | OCB | CSF [IgG] | IgG Index | Alb Index | Previously Sequenced |
| --- | --- | --- | --- | --- | --- | --- | --- | --- |
| **DD-03** | 30's | M | ADEM | Positive 3 Bands | **30.0** | **0.85** | **31.8** | Yes |
| **DD-08** | 30's | F | RRMS | Positive 5 Bands | 1.0 | 0.55 | 6.0 | No |
| **DD-10** | 40's | F | Neurodegeneration, MS | Positive, 3 Bands | 2.0 | **0.89** | 1.4 | No |
| **DD-11** | 20's | F | Tumefactive MS | Negative | 0.9 | ND | ND | No |
| **DD-13** | 60's | M | ADEM | Negative | **6.1** | **0.72** | **12.3** | No |
| **DD-17** | 50's | F | Tumefactive MS | Negative Matched | 1.8 | ND | ND | Yes |
| **DD-19** | 30's | F | Untyped MS | Negative 1 Band | 1.9 | ND | ND | Yes |
| **DD-21** | 60's | F | Untyped MS | Negative | 1.5 | ND | ND | Yes |
| **DD-71** | 30's | F | Untyped MS | Positive 6 Bands | **7.7** | **0.71** | **15.5** | No |
| **DD-72** | 50's | M | RRMS | Negative Matched | 2.5 | 0.49 | 6.0 | Yes |
| **DD-79** | 50's | M | Rhombencephalitis, ADEM | Positive 4 Bands | 3.6 | **0.88** | **11.0** | No |
| **DD-80** | 40's | M | ADEM | Negative | **11.6** | **0.78** | **14.8** | No |
| **DD-82** | 20's | F | NMOSD | Negative Matched | 4.0 | 0.49 | 6.9 | No |
| **DD-83** | 20's | F | Tumefactive MS | Positive 6 Bands | 2.0 | **0.75** | 3.4 | No |
| **OND-63** | teens | F | Migraine, Spina Bifida | ND | <0.3 | ND | ND | No |
| **OND-64** | 30's | F | Microvascular Disease | Negative Matched | 3.1 | 0.38 | 5.3 | No |
| **OND-73** | 60's | M | Atypical Stroke | Negative | 5.6 | **0.72** | **10.5** | No |
| **OND-75** | 40's | F | CSF Shunt Infection | ND | ND | ND | ND | No |
| **OND-76** | 30's | M | Pudendal Neuralgia | Negative | 2.3 | 0.47 | 7.8 | No |
| **OND-78** | 40's | M | CNS Vasculitis | Negative | 2.6 | 0.54 | 8.0 | No |
| **OND-81** | 70's | F | CNS Lyme Disease | Positive 13 Bands | **9.1** | **1.10** | **10.3** | No |
| **OND-84** | 50's | M | Neurosyphilis | Negative | **10.3** | 0.55 | **10.4** | No |
| **C-23** | 70's | M | NPH | ND | <0.3 | ND | ND | No |
| **C-25** | 30's | F | IIH |  | 1.1 |  |  |  |
| **C-27** | 70's | M | NPH |  | 0.9 |  |  |  |
| **C-28** | 30's | F | IIH |  | <0.3 |  |  |  |
| **C-30** | 60's | F | NPH |  | 0.9 |  |  |  |
| **C-31** | 60's | M | NPH |  | <0.3 |  |  |  |
| **C-32** | 30's | M | Cranioplasty, Stroke |  | 2.5 |  |  |  |
| **C-33** | 50's | F | Acute Hydrocephalus |  | <0.3 |  |  |  |
| **C-34** | 70's | M | NPH |  | 1.2 |  |  |  |
| **C-58** | 30's | M | Hydrocephalus |  | <0.3 |  |  |  |
| **C-59** | 60's | M | Hydrocephalus |  | 1.0 |  |  |  |
| **C-67** | 70's | M | NPH |  | 1.9 |  |  |  |
| C-68 | 20's | M | Hydrocephalus |  | <0.3 |  |  |  |

Age is provided as decade of life (e.g. “40’s”) per MedRxiv requirement
CSF [IgG] = concentration of IgG in CSF (normal 0.0 – 6.0 mg/dl), elevated values in **bold**
IgG Index = ratio of the quotients for IgG and albumin ( $[\text{IgG}]_{\text{csf}}/\text{IgG}_s)/([\text{Alb}]_{\text{csf}}/[\text{Alb}]_s)$ , (normal 0.28 – 0.66) , elevated values in **bold**
Alb Index = Albumin Index (normal 0.0 – 9.0), elevated values in **bold**
DD = demyelinating disease subject
OND = other neurologic disease subject
C = control subject
OCB = oligoclonal band testing
ADEM = acute disseminated encephalomyelitis

### Clinical findings in the MS and OND groups

Six of the 14 DD subjects had positive OCB testing (Table 1; DD-03, 08, 10, 71, 79, 83). Two of the DD patients who had negative OCB tests had CSF collected within 3 days of their disease onset (DD-80, 82), while 2 others had matched bands in serum and CSF (DD-17, 72) and one had only a single band in CSF. It is possible that some of these subjects might have developed OCBs later (e.g. DD-80, 82), but, unfortunately, CSF was not recollected in these subjects after their initial hospitalizations and enrollment in the study. Three subjects had CSF collected after many years of stable disease (DD-17, 19, 21). Six of the eight OND subjects also had OCB testing. Among these one was positive, which was a subject with proven CNS Lyme disease (OND-81).

Ten of the 14 DD subjects had albumin index determinations, a measure of blood-brain barrier (BBB) intactness. Five of 10 albumin index determinations were normal (< 9.0), providing evidence for an intact BBB in half the DD subjects.

### Spinal fluid serologic testing against MS candidate bacteria

All the DD, OND, and control specimens were tested for the CSF IgG responses to each organism, expressed as ELISA Index (EI). This data is displayed in Figure 1 and tabulated in Tables 3 and 4.

**Table 3.** Summary of CSF serologic responses to the 10 MS candidate bacteria. Indirect ELISA was performed on CSF from 14 subjects with definite demyelinating disease. Commercial human CSF (Randox Laboratories Ltd, UK; ([IgG] 10 mg/dl) was used as the positive control. IgGdepleted commercial human CSF (Randox Laboratories Ltd, UK) was utilized as the negative (calibration) control. The EI of the negative control is defined as 1.0.

| Subject | Neurologic Diagnosis | OCB | BBB Intact | Akk | Atop | Bact | Lacto | Odor | Pseudo | Cuti | Fuso | Porphy | Strep |
| --- | --- | --- | --- | --- | --- | --- | --- | --- | --- | --- | --- | --- | --- |
| DD-03 | ADEM | Positive<br>3 Bands | No | 10.6 | 12.1 | 5.2 | 11.6 | 1.9 | 8.6 | 4.2 | 0.6 | 2.9 | 2.7 |
| DD-08 | RRMS | Positive<br>5 Bands | Yes | 2.3 | 3.7 | 1.8 | 1.6 | 1.8 | 2.6 | 6.2 | qns | qns | qns |
| DD-10 | Progressive MS | Positive<br>3 Bands | Yes | 5.4 | 5.3 | 2.2 | 1.8 | 1.9 | 4.3 | 6.3 | 1.6 | 2.0 | 2.9 |
| DD-11 | MS untyped | Negative | ND | 1.5 | 2.2 | 1.3 | 1.3 | 1.7 | 2.0 | qns | qns | qns | qns |
| DD-13 | ADEM | Negative | No | 3.7 | 6.8 | 1.8 | 5.0 | 1.8 | 9.2 | qns | qns | qns | qns |
| DD-17 | Tumefactive MS | Negative Matched | ND | 3.3 | 4.5 | 1.2 | 1.3 | 1.2 | 2.4 | 4.3 | 1.0 | 0.9 | 1.8 |
| DD-19 | MS Untyped | Negative<br>1 Band | ND | 2.2 | 1.6 | 1.0 | 3.1 | 2.8 | 1.2 | 6.7 | 1.0 | 0.9 | 1.9 |
| DD-21 | CIS | Negative | ND | 4.9 | 4.2 | 2.4 | 3.1 | 4.7 | 3.1 | 5.2 | 2.0 | 2.4 | 1.8 |
| DD-71 | RRMS | Positive<br>6 Bands | No | 4.7 | 7.3 | 3.1 | 6.2 | 2.7 | 4.8 | 3.8 | 1.4 | 1.7 | 0.6 |
| DD-72 | RRMS | Negative<br>Matched | Yes | 7.1 | 4.3 | 2.6 | 4.1 | 5.8 | 3.8 | 5.3 | 1.3 | 1.4 | 2.9 |
| DD-79 | Rhombencephalitis | Positive<br>4 Bands | No | 1.7 | 3.8 | 1.3 | 1.7 | 1.5 | 1.8 | 5.6 | 1.1 | 1.2 | 2.2 |
| DD-80 | Tumefactive MS | Negative | No | 2.2 | 3.7 | 2.8 | 8.2 | 2.4 | 2.3 | 6.6 | 1.0 | 1.4 | 2.7 |
| DD-82 | NMOSD | Negative Matched | Yes | 3.4 | 4.7 | 3.1 | 10.4 | 4.7 | 3.8 | 6.7 | 1.0 | 1.6 | 3.1 |
| DD-83 | Tumefactive MS | Positive<br>6 Bands | Yes | 2.2 | 2.1 | 2.3 | 3.8 | 3.4 | 1.5 | 5.6 | 0.8 | 1.4 | 2.0 |
| Positive Control | - | - | - | 6.6 | 9.2 | 5.2 | 9.3 | 5.4 | 7.3 | 4.0 | 1.9 | 3.4 | 4.1 |

EI = ELISA Index Value: $EI \leq 1.0$ = negative $EI 1.1 - 2.9$ = weak positive $EI 3.0 - 4.9$ = positive $EI \geq 5.0$ = strong positive
DD = demyelinating disease, CIS = clinically isolated syndrome, ADEM = acute disseminated encephalomyelitis,
RRMS = relapsing-remitting multiple sclerosis, OCB = results of clinical oligoclonal band testing,
BBB = blood-brain barrier intactness, based on the albumin index value where normal (0-9) is intact, > 9 is compromised,
ND = not tested qns = quantity not sufficient for testing
NMOSD = NMO spectrum disorder
Akk = *Akkermansia muciniphila*; Lacto = *Lactobacillus paracasei*; Pseudo = *Pseudomonas aeruginosa*; Atop = *Atopobium vaginae*; Bact = *Bacteroides fragilis*; Odor = *Odoribacter splanchnicus*; Strep = *Streptococcus mutans*; Cuti = *Cutibacterium acnes*; Porphy = *Porphyromonas gingivalis*; Fuso = *Fusobacterium necrophorum*

**Table 4.**
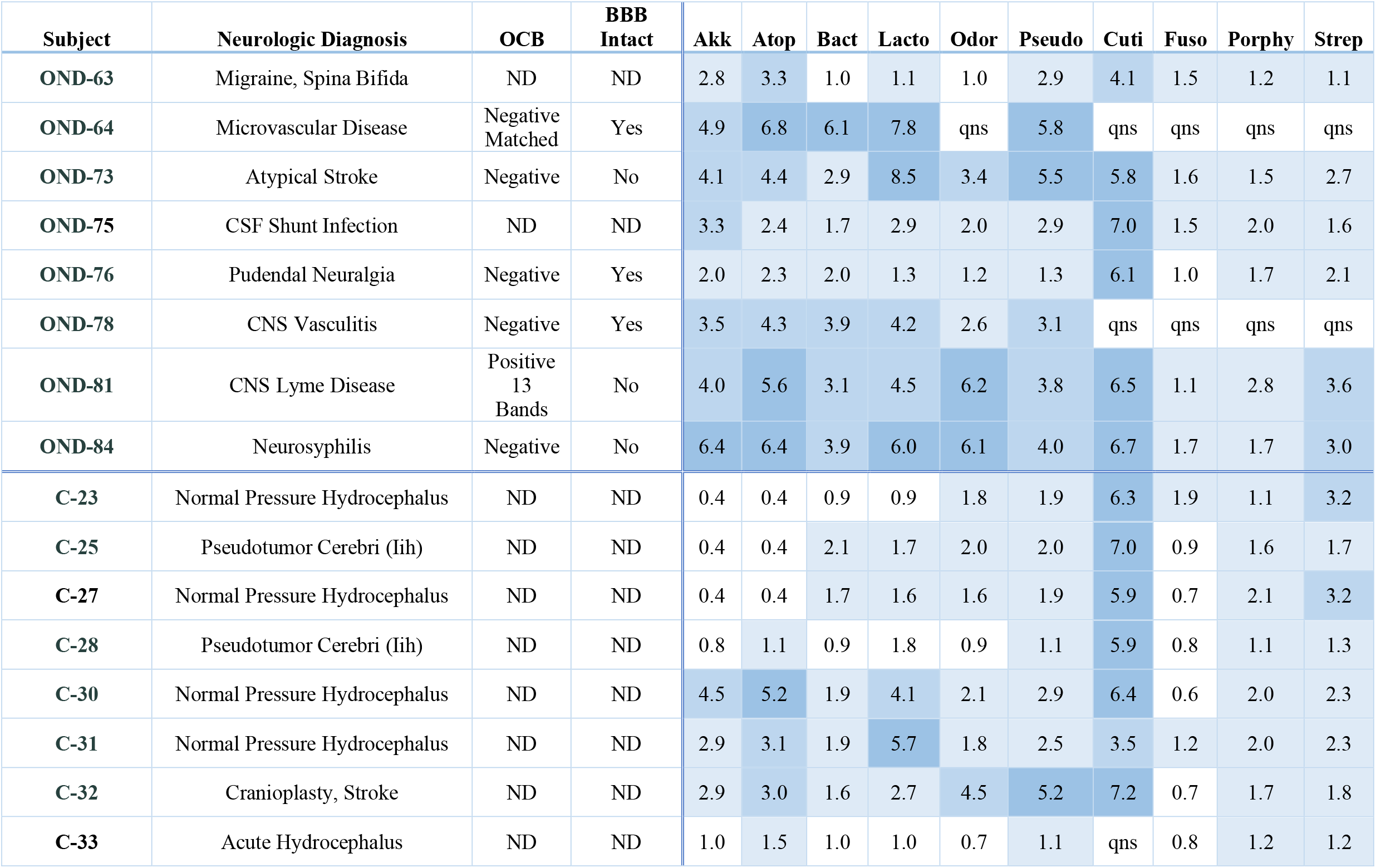
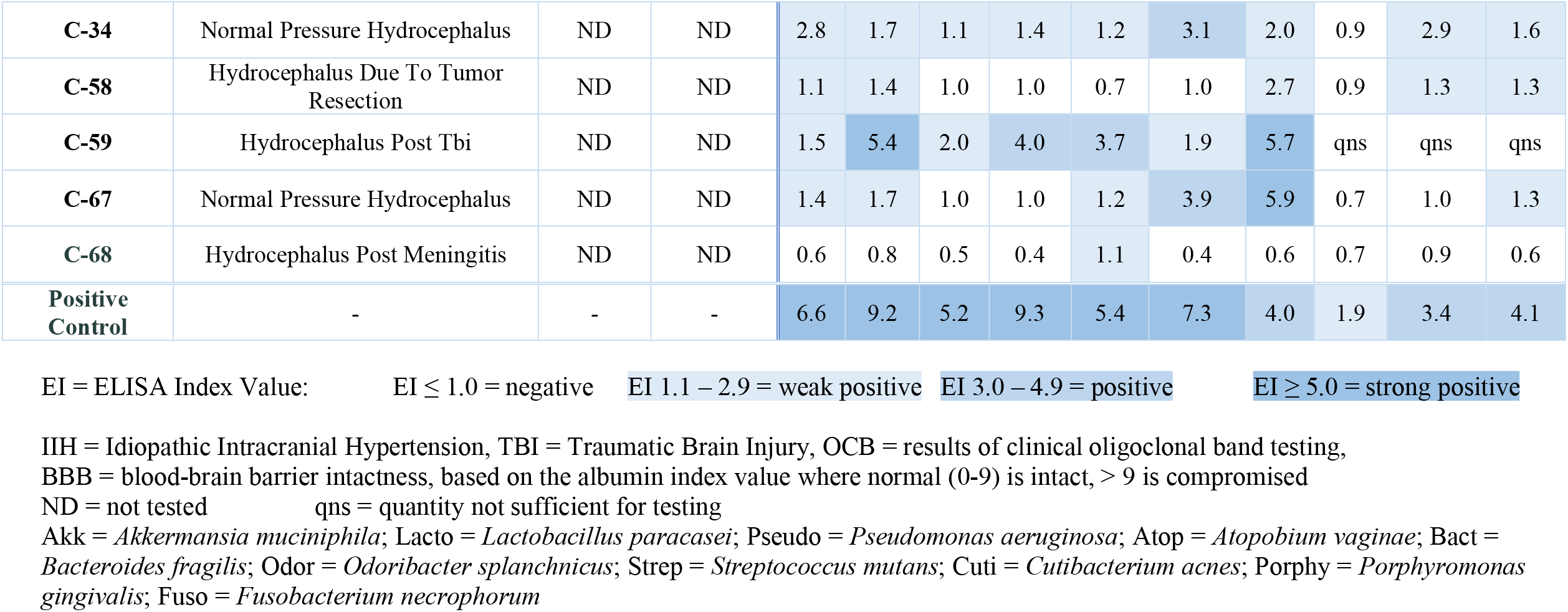
Summary of CSF serologic responses to the MS candidate bacteria. Indirect ELISA was performed on CSF from 8 subjects with other neurologic diseases (OND) and on CSF from 13 control subjects. Commercial human CSF (Randox Laboratories Ltd, UK; ([IgG] 10 mg/dl) was used as the positive control. IgG-depleted commercial human CSF (Randox Laboratories Ltd, UK) was utilized as the negative (calibration) control. The EI of the negative control is defined as 1.0.

| Subject | Neurologic Diagnosis | OCB | BBB Intact | Akk | Atop | Bact | Lacto | Odor | Pseudo | Cuti | Fuso | Porphy | Strep |
| --- | --- | --- | --- | --- | --- | --- | --- | --- | --- | --- | --- | --- | --- |
| OND-63 | Migraine, Spina Bifida | ND | ND | 2.8 | 3.3 | 1.0 | 1.1 | 1.0 | 2.9 | 4.1 | 1.5 | 1.2 | 1.1 |
| OND-64 | Microvascular Disease | Negative Matched | Yes | 4.9 | 6.8 | 6.1 | 7.8 | qns | 5.8 | qns | qns | qns | qns |
| OND-73 | Atypical Stroke | Negative | No | 4.1 | 4.4 | 2.9 | 8.5 | 3.4 | 5.5 | 5.8 | 1.6 | 1.5 | 2.7 |
| OND-75 | CSF Shunt Infection | ND | ND | 3.3 | 2.4 | 1.7 | 2.9 | 2.0 | 2.9 | 7.0 | 1.5 | 2.0 | 1.6 |
| OND-76 | Pudendal Neuralgia | Negative | Yes | 2.0 | 2.3 | 2.0 | 1.3 | 1.2 | 1.3 | 6.1 | 1.0 | 1.7 | 2.1 |
| OND-78 | CNS Vasculitis | Negative | Yes | 3.5 | 4.3 | 3.9 | 4.2 | 2.6 | 3.1 | qns | qns | qns | qns |
| OND-81 | CNS Lyme Disease | Positive 13 Bands | No | 4.0 | 5.6 | 3.1 | 4.5 | 6.2 | 3.8 | 6.5 | 1.1 | 2.8 | 3.6 |
| OND-84 | Neurosyphilis | Negative | No | 6.4 | 6.4 | 3.9 | 6.0 | 6.1 | 4.0 | 6.7 | 1.7 | 1.7 | 3.0 |
| C-23 | Normal Pressure Hydrocephalus | ND | ND | 0.4 | 0.4 | 0.9 | 0.9 | 1.8 | 1.9 | 6.3 | 1.9 | 1.1 | 3.2 |
| C-25 | Pseudotumor Cerebri (Iih) | ND | ND | 0.4 | 0.4 | 2.1 | 1.7 | 2.0 | 2.0 | 7.0 | 0.9 | 1.6 | 1.7 |
| C-27 | Normal Pressure Hydrocephalus | ND | ND | 0.4 | 0.4 | 1.7 | 1.6 | 1.6 | 1.9 | 5.9 | 0.7 | 2.1 | 3.2 |
| C-28 | Pseudotumor Cerebri (Iih) | ND | ND | 0.8 | 1.1 | 0.9 | 1.8 | 0.9 | 1.1 | 5.9 | 0.8 | 1.1 | 1.3 |
| C-30 | Normal Pressure Hydrocephalus | ND | ND | 4.5 | 5.2 | 1.9 | 4.1 | 2.1 | 2.9 | 6.4 | 0.6 | 2.0 | 2.3 |
| C-31 | Normal Pressure Hydrocephalus | ND | ND | 2.9 | 3.1 | 1.9 | 5.7 | 1.8 | 2.5 | 3.5 | 1.2 | 2.0 | 2.3 |
| C-32 | Cranioplasty, Stroke | ND | ND | 2.9 | 3.0 | 1.6 | 2.7 | 4.5 | 5.2 | 7.2 | 0.7 | 1.7 | 1.8 |
| C-33 | Acute Hydrocephalus | ND | ND | 1.0 | 1.5 | 1.0 | 1.0 | 0.7 | 1.1 | qns | 0.8 | 1.2 | 1.2 |
| **C-34** | Normal Pressure Hydrocephalus | ND | ND | 2.8 | 1.7 | 1.1 | 1.4 | 1.2 | 3.1 | 2.0 | 0.9 | 2.9 | 1.6 |
| **C-58** | Hydrocephalus Due To Tumor Resection | ND | ND | 1.1 | 1.4 | 1.0 | 1.0 | 0.7 | 1.0 | 2.7 | 0.9 | 1.3 | 1.3 |
| **C-59** | Hydrocephalus Post Tbi | ND | ND | 1.5 | 5.4 | 2.0 | 4.0 | 3.7 | 1.9 | 5.7 | qns | qns | qns |
| **C-67** | Normal Pressure Hydrocephalus | ND | ND | 1.4 | 1.7 | 1.0 | 1.0 | 1.2 | 3.9 | 5.9 | 0.7 | 1.0 | 1.3 |
| **C-68** | Hydrocephalus Post Meningitis | ND | ND | 0.6 | 0.8 | 0.5 | 0.4 | 1.1 | 0.4 | 0.6 | 0.7 | 0.9 | 0.6 |
| **Positive Control** | - | - | - | 6.6 | 9.2 | 5.2 | 9.3 | 5.4 | 7.3 | 4.0 | 1.9 | 3.4 | 4.1 |

EI = ELISA Index Value: EI ≤ 1.0 = negative EI 1.1 – 2.9 = weak positive EI 3.0 – 4.9 = positive EI ≥ 5.0 = strong positive
IIH = Idiopathic Intracranial Hypertension, TBI = Traumatic Brain Injury, OCB = results of clinical oligoclonal band testing,
BBB = blood-brain barrier intactness, based on the albumin index value where normal (0-9) is intact, > 9 is compromised
ND = not tested qns = quantity not sufficient for testing
Akk = *Akkermansia muciniphila*; Lacto = *Lactobacillus paracasei*; Pseudo = *Pseudomonas aeruginosa*; Atop = *Atopobium vaginae*; Bact = *Bacteroides fragilis*; Odor = *Odoribacter splanchnicus*; Strep = *Streptococcus mutans*; Cuti = *Cutibacterium acnes*; Porphy = *Porphyromonas gingivalis*; Fuso = *Fusobacterium necrophorum*

**Figure 1.**
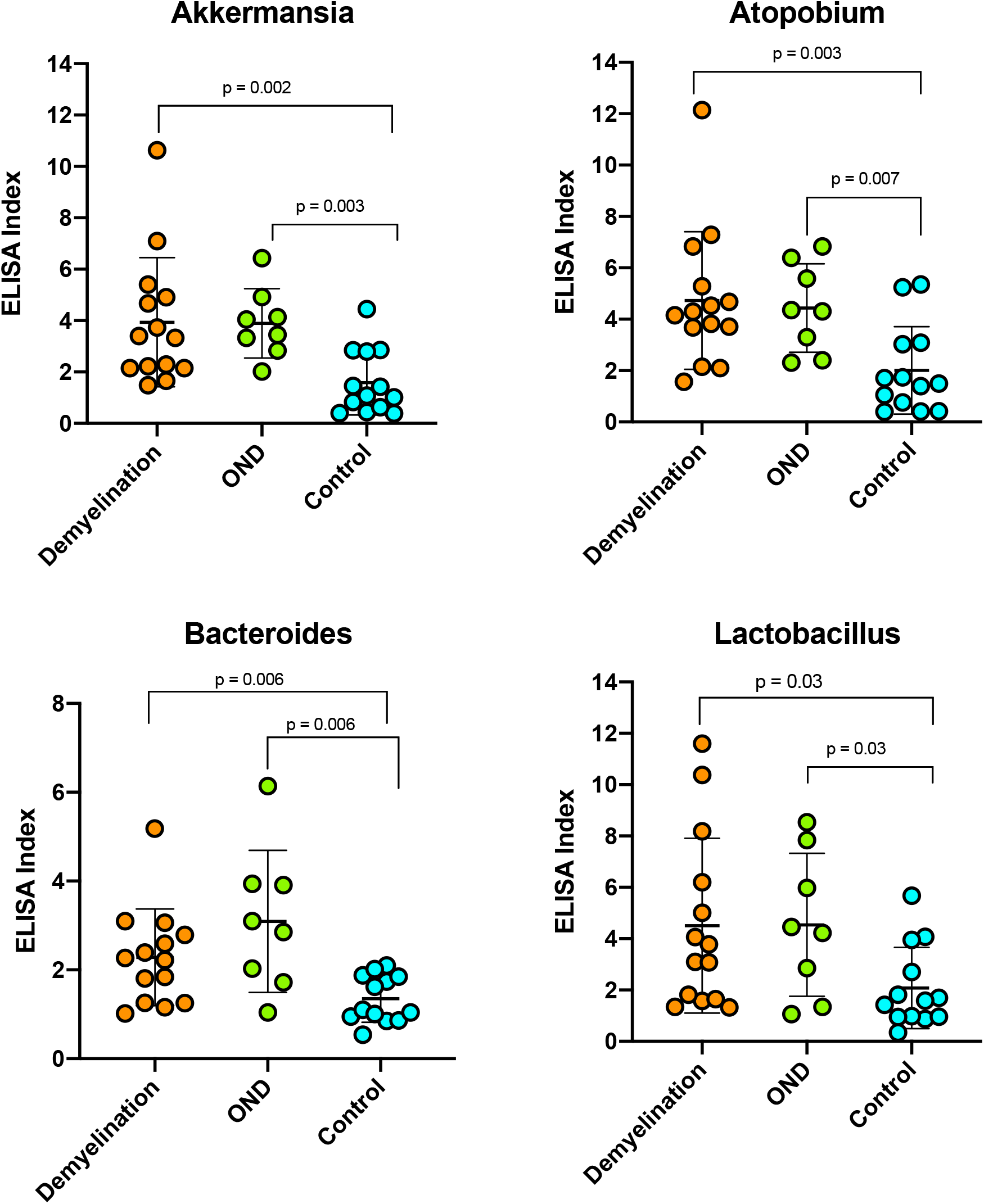
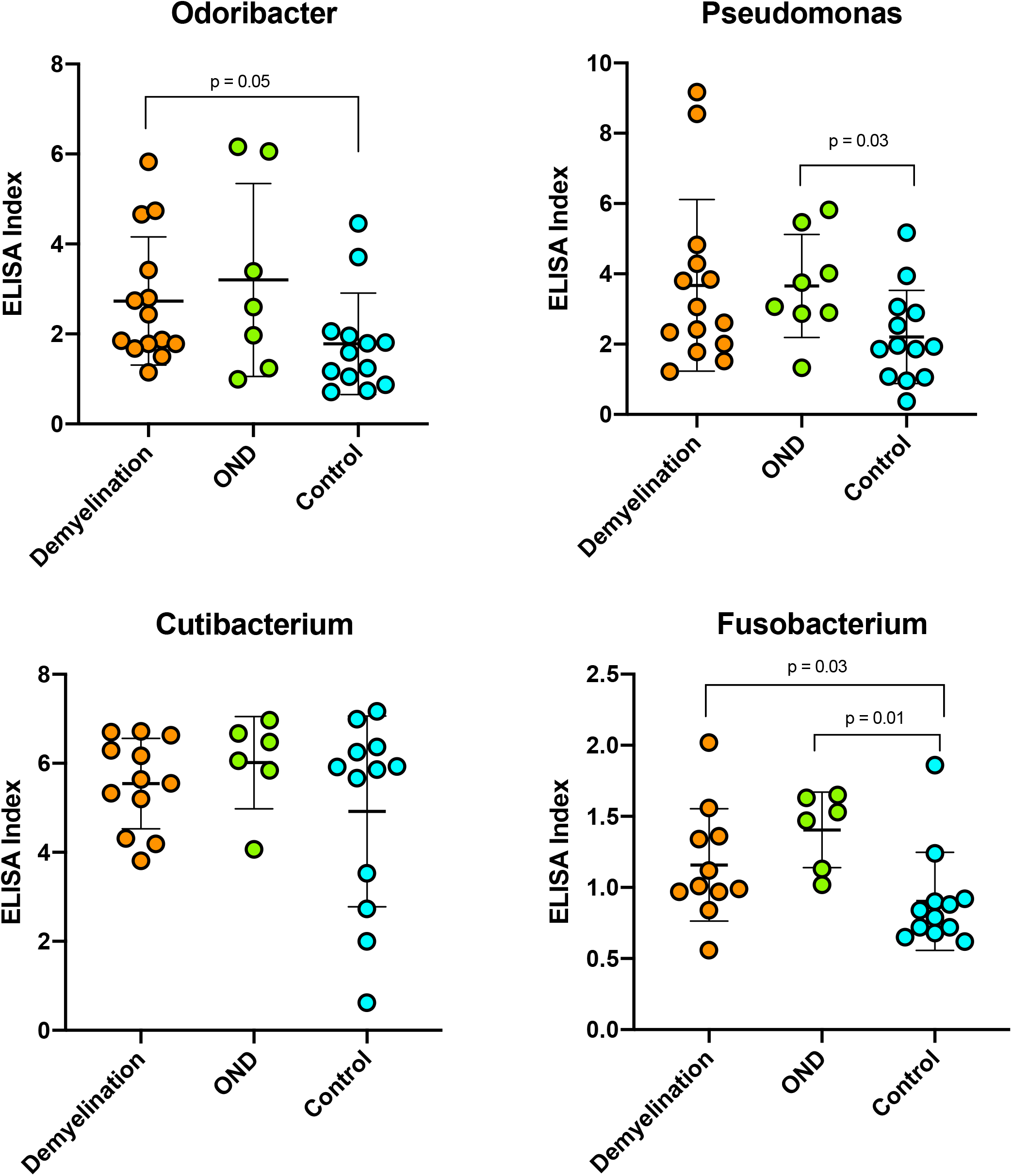
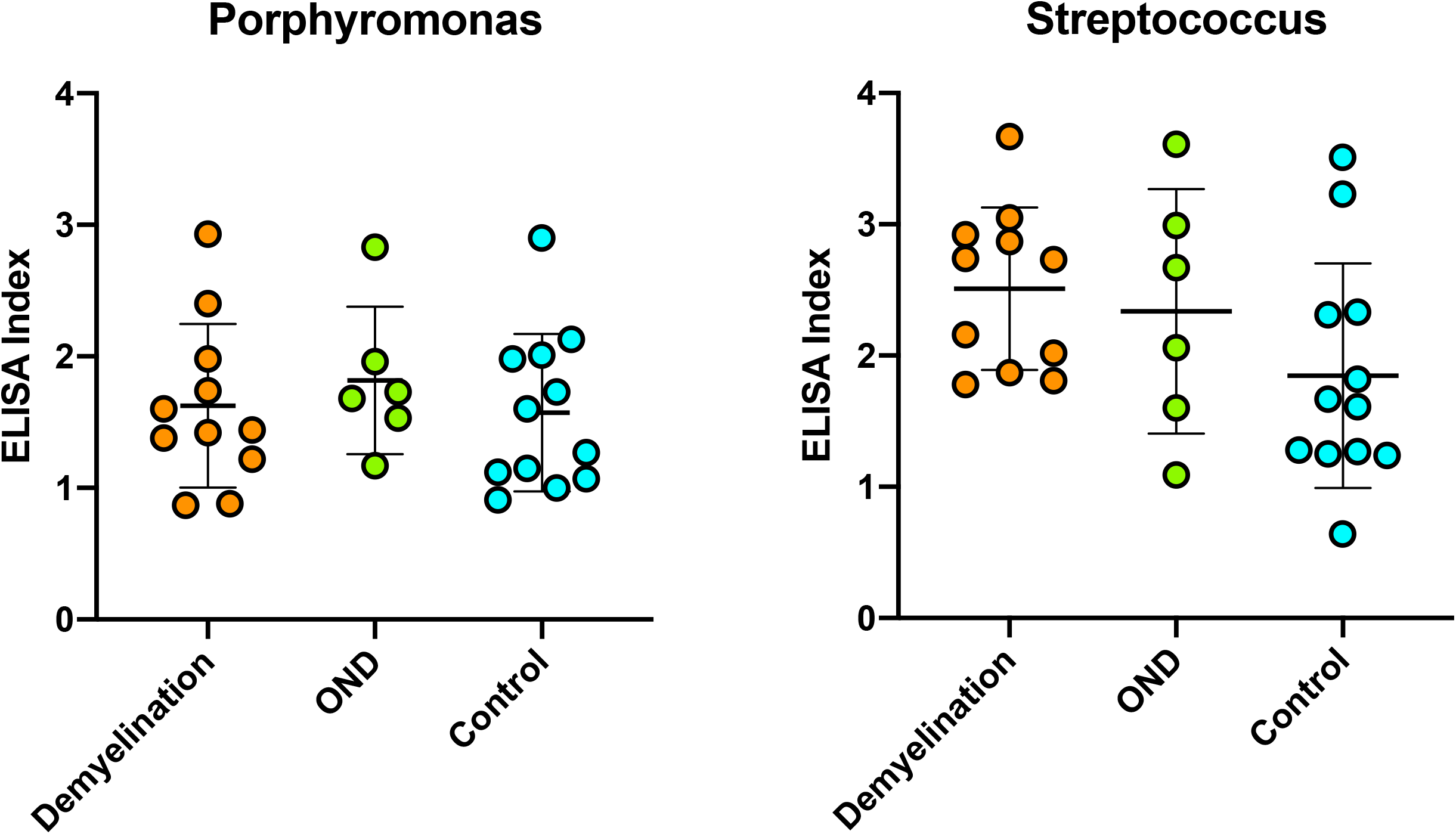
ELISA Index values for each of the 10 MS candidate bacteria. Indirect ELISA was performed on CSF from subjects with definite demyelinating disease, other neurologic diseases (OND), and controls. Statistical comparisons were made using the Mann-Whitney nonparametric test. The bars indicate mean values ± standard deviation. See Table S2 for additional information.

**Figure 2.**
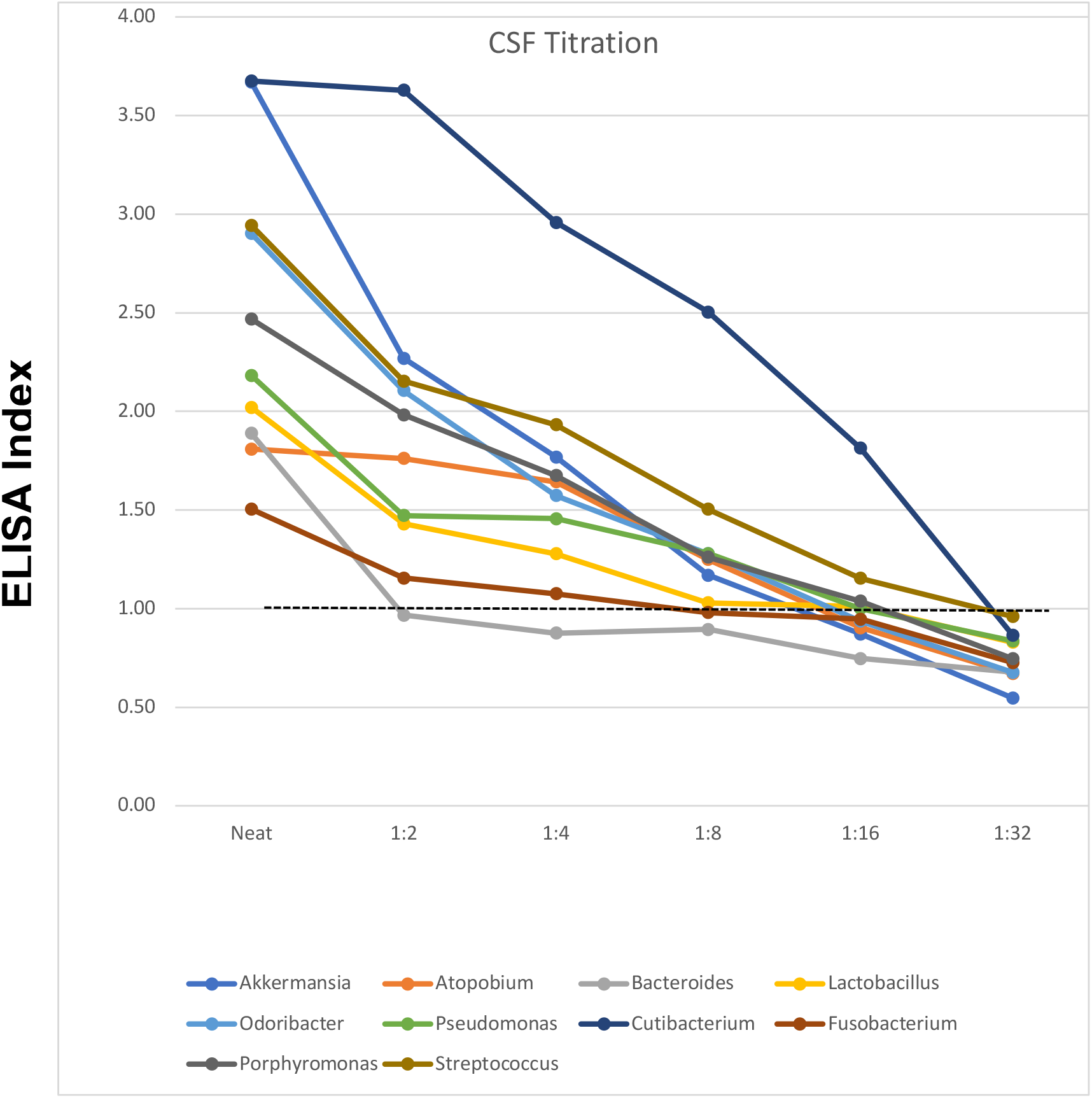
ELISA index following CSF dilution. CSF was serially diluted 1:2 in phosphate buffered saline and subjected to indirect ELISA against each of the 10 antigens. CSF was selected from subjects with previous reactivity against the selected antigen. ELISA index values are plotted against the 2-fold dilutions. The calibration control (Randox CSF, IgG depleted) for each antigen has an EI value of 1.0 by definition (dashed line). In this manner, one can establish end-point dilutions (where EI > 1.0) for each sample against each antigen. For instance, the endpoint dilution for Cutibacterium (dark blue) is 1:16, and for Fusobacterium (brown) 1:4. CSF specimens were taken from subjects: DD-72 – Akkermansia, Odoribacter, Streptococcus; DD-71 – Atopobium; C-25 – Bacteroides; C-31 – Lactobacillus; C-32 – Pseudomonas; DD-21 – Fusobacterium, Porphyromonas

Measured CSF seroreactivity (mean EI) was greatest against Cutibacterium, Atopobium, and Lactobacillus, and lowest against Fusobacterium and Porphyromonas (Tables 3, 4, and S2). There were significant differences in CSF reactivity among the groups against Akkermansia, Atopobium, Bacteroides, Lactobacillus, and Fusobacterium. CSF seroreactivity was similar between the DD and OND groups. Reactivity was significantly higher in the DD group compared to the controls against Akkermansia, Atopobium, Bacteroides, Lactobacillus, Odoribacter, and Fusobacterium (Figure 1, Table S2). Reactivity was significantly higher in the OND group compared to the controls against Akkermansia, Atopobium, Bacteroides, Lactobacillus, and Pseudomonas.

Among the 129 CSF serologic tests reported from the DD group, 25 (19%) were strongly positive (EI ≥ 5.0), 29 (22%) were positive (EI 3.0-4.9), 65 (50%) were equivocal (EI 1.1-2.9), and 10 (8%) were negative (EI ≤1.0). Among the 71 CSF serologic tests reported from the OND group, 17 (24%) were strongly positive, 20 (28%) were positive, 31 (50%) were equivocal, and 3 (4%) were negative (EI ≤1.0). Among the 126 CSF serologic tests reported from the Control group, 12 (10%) were strongly positive, 12 (10%) were positive, 61 (48%) were equivocal, and 41 (33%) were negative.

Within the DD group, subject DD-03, diagnosed with ADEM, had measurable reactivity in the CSF against 9 of the 10 bacterial antigens, and was strongly reactive against 5 antigens. The other ADEM subject, DD-13, displayed some reactivity against all 6 antigens tested, with strong reactivity against 3 of the 6.

#### Detection of Intrathecal Antibody Synthesis

Antibody indexes (AIs) were determined in 11 subjects from the DD group and 4 subjects from the OND group (Table 5). (Serum was not generally not collected from the control subjects, precluding calculation of AIs in this group.) Four of the 11 tested DD group subjects had Ais >1.0 against at least one of the 10 antigens, suggesting intrathecal production of anti-bacterial antibodies.^11^ One subject, DD-82 with NMO spectrum disorder had elevated AIs against 5 of the 10 antigens. Elevated AIs within the DD group were only modestly increased, ranging from 1-15% above the matched serum EI determinations. CSF from subject OND-78, a man with CNS vasculitis and normal albumin ration, had a much higher AI (3.61) measured against Odoribacter. The assay passed quality control measures, and duplicate determinations within the assay were similar. Unfortunately, this CSF specimen was limited so this value could not be confirmed with retesting.

**Table 5.**
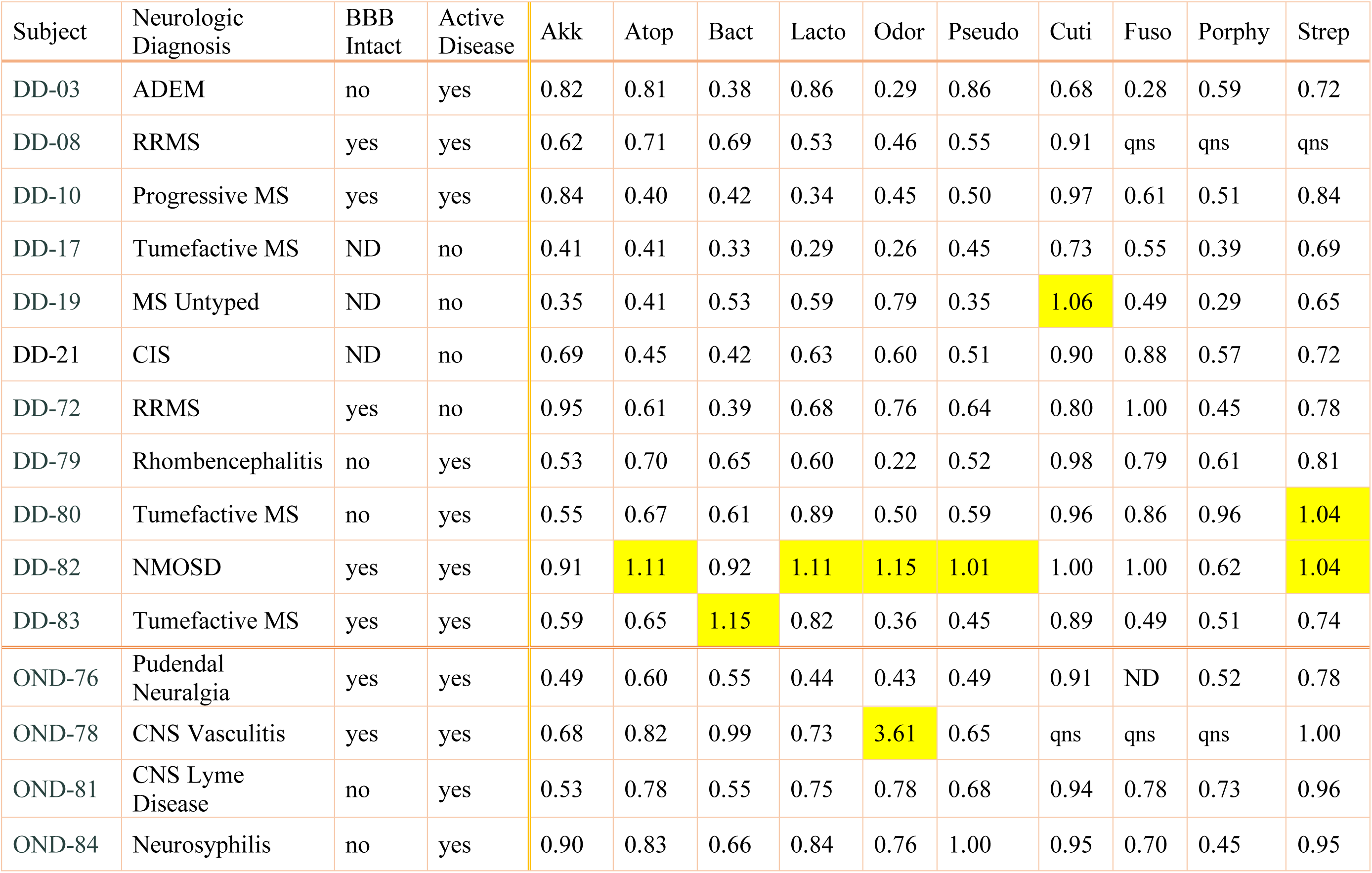
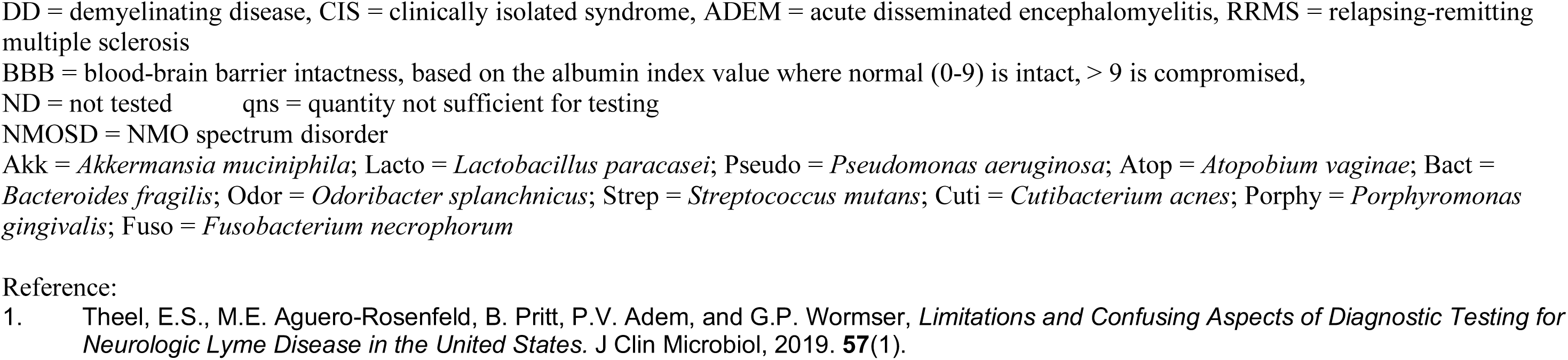
Summary of CSF:serum antibody indexes against the MS candidate bacteria. Serum samples were diluted in PBS to match the corresponding CSF IgG concentration. Then indirect ELISA was performed on CSF and diluted serum from the subjects. Each specimen was run in duplicate with CSF and diluted serum specimens on the same plate. Responses against each of the bacterial antigens were determined separately. Positive and negative controls were included with each assay. Antibody index (AI) values are defined as (CSF ELISA Index)/(Diluted Serum ELISA Index). AI values greater than 1.0 (highlighted) are considered evidence for the intrathecal production of antibody.1 AI could not be determined for some of the subjects (DD-11,13,17; OND-63,64,73,75) because serum was not collected.

The data allowed comparison of the measured EI’s with BBB intactness (albumin index) for each of the subjects (Figure S1). This is considered to be an exploratory analysis that, to our knowledge, has not been attempted before. Positive slopes (EI/AI) were observed for 5 of the 10 tested antigens, suggesting that higher anti-bacterial Ab responses correlated with less intact BBB (higher albumin index). Data points above the expected EI (line) for a given albumin index (e.g. subjects DD-10 and DD-72 in the anti-Akkermansia plot) are interpreted as evidence for intrathecal antibody synthesis. Subject DD-82 had higher than expected EIs against Bacteroides, Lactobacillus, and Odoribacter (Figure S1), partially corresponding to elevated AIs against Atopobium, Lactobacillus, Odoribacter, Pseudomonas, and Streptococcus (Table 5).

#### Comparison with the Updated Sequencing Findings

DD subjects 03, 17, 19, 21, and 72 all had surgically obtained brain tissue sequenced and analyzed using the methods previously described.^6^ Updated sequencing results are reported in Table S3. Comparisons of EI values and sequencing results are shown in Table 6. The intervals between brain biopsy and CSF collections were highly variable, ranging from 1 week to 10 years. Many but not all of the CSF EI values are supported by sequencing reads. For instance, in subject DD-03 the most abundant sequence mappings are to Lactobacillus, Streptococcus, and Cutibacterium. CSF EI values are highest for this subject against Lactobacillus and Cutibacterium, with less reactivity against Streptococcus. All the 5 sequenced subjects’ CSF samples were reactive against Akkermansia, and there were at least a few sequencing reads mapping to this bacterium among all 5 subjects. In contrast, all 5 subjects’ CSF samples were also reactive against Odoribacter, but only 2 of 5 corresponding brain tissue samples had reads that mapped to this bacterium. These apparent discrepancies may be ascribed to the sometimes prolonged intervals between the brain biopsies and CSF collections, or to cross reactivity to other bacteria among the ELISA assays.

**Table 6.**
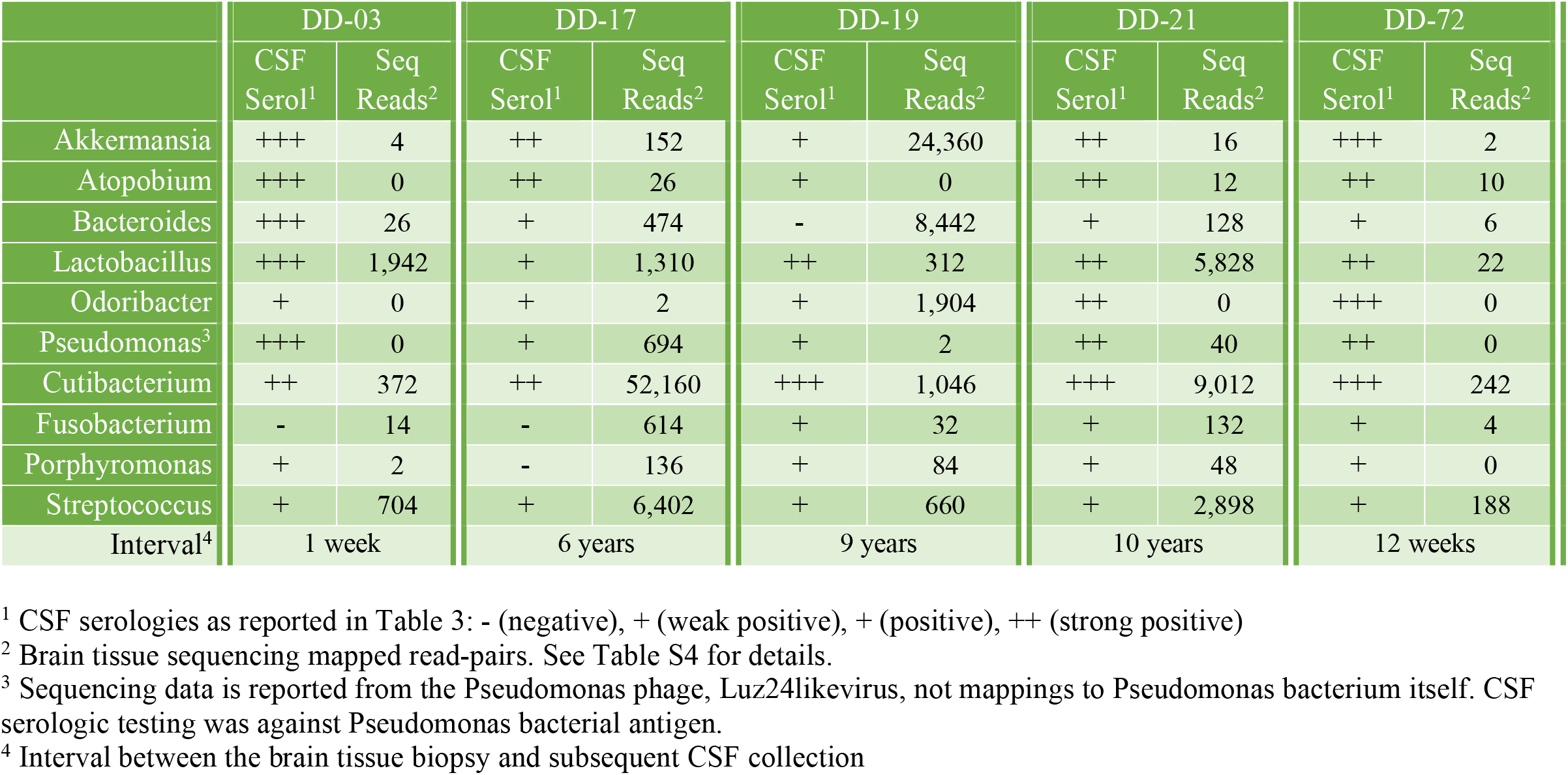
Comparison of CSF serology and brain tissue sequencing results. Subjects DD-03, DD-17, DD-19, DD-21, and DD-72 had both brain tissue sequencing and CSF serology performed. The interval between the diagnostic brain biopsy and subsequent CSF collection was variable.

|  | DD-03 |  | DD-17 |  | DD-19 |  | DD-21 |  | DD-72 |  |
| --- | --- | --- | --- | --- | --- | --- | --- | --- | --- | --- |
|  | CSF Serol<sup>1</sup> | Seq Reads<sup>2</sup> | CSF Serol<sup>1</sup> | Seq Reads<sup>2</sup> | CSF Serol<sup>1</sup> | Seq Reads<sup>2</sup> | CSF Serol<sup>1</sup> | Seq Reads<sup>2</sup> | CSF Serol<sup>1</sup> | Seq Reads<sup>2</sup> |
| Akkermansia | +++ | 4 | ++ | 152 | + | 24,360 | ++ | 16 | +++ | 2 |
| Atopobium | +++ | 0 | ++ | 26 | + | 0 | ++ | 12 | ++ | 10 |
| Bacteroides | +++ | 26 | + | 474 | - | 8,442 | + | 128 | + | 6 |
| Lactobacillus | +++ | 1,942 | + | 1,310 | ++ | 312 | ++ | 5,828 | ++ | 22 |
| Odoribacter | + | 0 | + | 2 | + | 1,904 | ++ | 0 | +++ | 0 |
| Pseudomonas<sup>3</sup> | +++ | 0 | + | 694 | + | 2 | ++ | 40 | ++ | 0 |
| Cutibacterium | ++ | 372 | ++ | 52,160 | +++ | 1,046 | +++ | 9,012 | +++ | 242 |
| Fusobacterium | - | 14 | - | 614 | + | 32 | + | 132 | + | 4 |
| Porphyromonas | + | 2 | - | 136 | + | 84 | + | 48 | + | 0 |
| Streptococcus | + | 704 | + | 6,402 | + | 660 | + | 2,898 | + | 188 |
| Interval<sup>4</sup> | 1 week |  | 6 years |  | 9 years |  | 10 years |  | 12 weeks |  |

<sup>1</sup> CSF serologies as reported in Table 3: - (negative), + (weak positive), + (positive), ++ (strong positive)
<sup>2</sup> Brain tissue sequencing mapped read-pairs. See Table S4 for details.
<sup>3</sup> Sequencing data is reported from the Pseudomonas phage, Luz24likevirus, not mappings to Pseudomonas bacterium itself. CSF serologic testing was against Pseudomonas bacterial antigen.
<sup>4</sup> Interval between the brain tissue biopsy and subsequent CSF collection

## DISCUSSION

We conducted a study of CSF reactivity against 10 MS microbial candidates previously identified by RNA sequencing of diseased brain tissue. CSF reactivity was significantly higher in the demyelination group compared to the controls against 6 of the bacterial antigens: Akkermansia, Atopobium, Bacteroides, Lactobacillus, Odoribacter, and Fusobacterium. Four of the 11 tested DD group subjects had elevated antibody indexes against at least one of the 10 antigens, suggesting at least limited intrathecal production of anti-bacterial antibodies. This study clearly shows that there are antibacterial IgG antibodies against several MS candidate bacteria in most of the CSF specimens tested in the DD and OND groups. The findings are consistent with the hypothesis that MS candidate microbes trigger demyelination in some of these patients.

The experiments and analysis were performed in the most straightforward manner possible – direct comparisons between undiluted CSF in all the groups, and serum dilutions matched exactly to CSF IgG concentrations for the determinations of AI. The authors understand that there will be some leakage of antibodies from serum into the CSF, and that this is part of the pathologic process that defines the disease. Blood-brain barrier dysfunction in MS is well established, also reflected by IgG access to the parenchyma, a parameter used by neuropathologists to identify sites with barrier breach. However, not all the subjects in this study had BBB dysfunction according to clinical measurement of the albumin index.

The current study does have some limitations. First, the reference specimens were taken from patients who had CSF shunts placed for hydrocephalus and related conditions. Therefore, the control specimens are not from completely healthy volunteers. Since CSF is not generally available from healthy donors, the uninfected shunt procedure samples were selected as the best reference group readily available.

Second, many of the MS and DD patients received some disease-modifying therapy (DMT). Therefore, we cannot exclude some drug effects on the anti-microbial responses in DD subjects, although most of the DD antibacterial EIs were higher than the controls, not lower as would be expected with immunosuppressive DMT.

Third, since it was not feasible to assess all 29 MS candidate microbes derived from the sequencing data, a rational and pragmatic selection of 10 major candidates was made.^6^ For instance, the top candidate identified from the sequencing data based on both mRNA load and prevalence, *Nitrosospira*, simply could not be cultivated in sufficient quantity to develop an ELISA, despite extensive efforts over several months.

Fourth, we focused this study on the detection of the human IgG isotype and its four subclasses. Recent elegant studies emphasize that gut-educated IgA B cells and plasma cells contribute to surveillance of the meningeal venous sinuses, and can limit CNS inflammation in EAE mouse models for MS.^15 16^ We did not assay for IgA or IgM in the CSF because IgG is the dominant form of immunoglobulin in human CSF with concentrations approximately 10-fold higher that IgA or IgM.^17^

Based on the commercial polyclonal and monoclonal positive control antibodies employed, the novel ELISAs demonstrated good specificity and sensitivity. Randox^®^ CSF was chosen as a reference for patient CSF because it is commercially available and relatively well characterized. This commercial CSF preparation containing 10 mg/dl IgG was used as a positive control for each of the assays. Randox^®^ CSF was also IgG-depleted for use as the negative or calibration control in each of the assays.^7-9^ This method was chosen as a reliable and reproducible method for determining a baseline level of CSF antibody reactivity between assays.

Within these ELISA assays we suspect there is considerable cross-reactivity, similar to that seen with the commercial polyclonal anti-*Pseudomonas aeruginosa* antibody used as a positive control in all the assays. Cross-reactivity between Lyme and syphilis testing is well recognized among clinicians who employ these anti-bacterial serologic tests in clinical practice.^18^ The results from the neurologic Lyme disease subject (OND-81) demonstrate cross-reactivity, where her CSF was strongly reactive against 3 of the MS bacterial antigens, and moderately reactive to 5 others. However, there was no elevation in AI for this subject against any of the 10 bacteria included in this study. *Borrelia burgdorferi* was not included as a candidate bacterium in this study, but clinical diagnostic testing showed clear intrathecal production of anti-*B. burgdorferi* antibodies. This patient did have an elevated albumin index suggesting a compromised BBB, so some anti-bacterial antibodies could have leaked into the CNS from the serum. Cross-reactivity could also be due to the commonly shared features of bacterial organisms such as the lipopolysaccharide (LPS), lipoteichoic acid (LTA), or peptidoglycan (PGN).

We argue that the data presented here supports a pathogenic role for several of the organisms in some of the subjects. That is, there is evidence of intrathecal antibody production against several of the MS candidate microbes in several of the subjects. Specifically, there were AI’s > 1.0, evidence for intrathecal antibacterial antibody synthesis, in DD subjects 19, 80, 82, and 83. Among these, subjects DD-19, 82, and 83 were relatively young women at the time of diagnosis, the most common demographic for the development of MS and related diseases. Comparisons of EI’s with the albumin index analysis (Figure S1) suggested that subjects DD-72 and DD-13 may also have intrathecal production of antibodies against some of the bacterial antigens tested. Elevated AI’s were observed against 5 bacterial antigens in subject DD-82, a young woman with neuromyelitis optica spectrum disorder. Interestingly, tumefactive MS developed in subject DD-83 about 3 weeks after brain surgery. She did not develop any signs of infection at the surgical site, and demyelination occurred both near this site and distant in the cerebellum. DD-83 had no evidence of MS prior to the demyelination event. AI was elevated only against Bacteroides, a gut anaerobe that is seldom associated with neurosurgical infections.

Previously, one of the current authors (J.D.L.) demonstrated anti-peptidoglycan antibodies in CSF collected from patients with active MS.^19^ This group also hypothesizes that bacterial cell wall peptidoglycan is involved in the development of CNS autoimmunity.^20, 21^ Another group has demonstrated that peptidoglycan contributes to demyelinating autoimmunity by engaging NOD1 and NOD2 in dendritic cells leading to downstream RIPK1 activation (a.k.a. RIP2 and RICK), promoting T-helper 17 (Th17) responses.^22^

Testing of CSF from subject DD-03 proved to be uniquely interesting. A possible inciting event was believed to be an attempted rescue of a newborn animal, possibly leading to a prolonged aerosol exposure within an enclosed space. This was followed by a brief febrile illness and then progressive neurologic dysfunction over the course of several weeks, with ADEM as the final neurologic diagnosis. Sequencing of a small diagnostic brain biopsy of this patient revealed lactobacillus as the most abundantly mapped microbe at the genus level. Lactobacillus is found in the female genital tract of many mammals, including humans, where it is dominant.^23, 24^ This subject’s CSF was highly reactive against 5 of the tested bacterial antigens, with the highest EI against *Lactobacillus paracasei* among all the subjects tested. However, the AI against lactobacillus (0.86) was not elevated in this subject, arguing against intrathecal antibody production. However, intrathecal antibody production is notoriously difficult to prove in individual subjects, methods vary, and a patient with compatible clinical findings should be considered to have the disease being considered, with or without elevated AI.^11, 25^ In sum, we argue that it is plausible that this subject’s ADEM was triggered by microbes, including *L. paracasei*, encountered from prolonged exposure to a newborn calf.

OCBs are a common clinical diagnostic feature of MS. For the DD cases CSF was taken about a week after DD-03s onset of neurologic symptoms, but DD-017, −019, and −021 had their CSF taken years after disease onset without any intervening attacks. DD-082 had her CSF collected only a few days after onset of disease, so it is possible that OCBs did not have time to develop. Additional CSF collections were not feasible in our population because, in general, the treating neurologists did not believe they were necessary or helpful for clinical care. It would be ideal to collect CSF at the time of attack, again a few months later, and around the time of any MS relapses, to assess if the antibacterial CSF responses become more focused over time.

Intrathecal production of antibodies against measles, rubella, and varicella zoster virus had been demonstrated in MS patients, although these antibodies account for less than 2% of total intrathecal IgG production.^26^ This MRZ reaction has been proposed as a relatively specific biomarker for MS.^27^ However, the prevailing view is that these polyspecific anti-viral IgGs do not normally correspond to the major oligoclonal IgG bands within the CSF and the MRZ reaction is considered to be a bystander reaction that does not reflect pathogenesis within the CNS. ^26, 28, 29^

The gut microbiota and CNS communicate bidirectionally, referred to as the gut-microbiota-brain axis and there are many pathways by which the two can interact, such as prolonged infection, molecular mimicry, bystander activation, and epitope spreading..^30-32^ Patients affected by MS exhibit a decrease in several gut microorganisms including *Bacteroides, Faecalibacterium*, and short-chain fatty acid (SCAFs) producing bacteria, with an increase in *Methanobrevibacter, Enterobacteriaceae*, and *Akkermansia*.^33, 34^ Treatment with disease-modifying therapies (DMTs) induce an increase of *Prevotella* compared with untreated patients. Moreover, colonization by *C. perfringens* type B is associated with relapse in MS patients, likely due to the toxins it produces, which can induce microvascular complications leading to neuronal and oligodendrocyte damage.^34-36^

In the CNS demyelinating disease, neuromyelitis optica (NMO), T-cell responses against aquaporin 4 (AQP4) p61-80 exhibit a Th17 bias, cross-reacting to *Clostridium perfringens* adenosine triphosphate-binding cassette transporter permease p204-217.^37^ Ninety percent amino acid homology was observed, which suggests that NMO autoimmunity may be driven by molecular mimicry at the T-cell level.

Similarly, for Guillain-Barré syndrome (GBS), serological studies demonstrated that most GBS patients who had anti-GBS antibodies also had anti-*Campylobacter jejuni* antibodies.^38^ GBS is an acute inflammatory polyradiculoneuropathy that typically develops following a gastrointestinal infection, with some reports suggesting a more severe form of the disease after *C. jejuni* infection.^39-42^ The cell wall of *C. jejuni* contains polysaccharides that resemble glycoconjugates of the human nerve tissues resulting in misguided attack on myelin and axons. Certain types of this disorder involve an immune response against gangliosides, which is suspected to originate due to molecular mimicry between gangliosides and lipopolysaccharides of *C. jejuni*.^43^

Due to the importance of the gut microbiome and immune function, Janji *et al*. assessed the gut microbiome within MS patients.^44^ They demonstrated that *Akkermansia muciniphila* was enriched in the stool of MS patients when compared to healthy controls and, when using a murine model for MS, altering the gut microbiome modulates CNS autoimmunity. A French group demonstrated anti-*Akkermansia muciniphila* immunoglobulin G (IgG) was increased in CSF from people with MS compared to controls, congruent with our findings.^45^ They also used indirect ELISA. However, this group did not find elevations in anti-*Fusobacterium necrophorum* or anti-*Bacteroides fragilis* antibodies in their CSF samples, while we did show an elevation in EIs in the DD group against these antigens.

In conclusion, the current study demonstrates that antibacterial antibodies can be detected in the CSF of subjects with demyelinating disease, and that CSF reactivity in the DD and OND groups exceeds reactivity in the controls for several bacterial species. This supports, but does not prove, the hypothesis that microbes contribute to demyelinating disease in persons with MS and related diseases with myelin loss. In the future, longitudinal studies investigating the possible intrathecal IgG and IgA production against different pathogens, development of sandwich ELISAs, and OCB subtraction are all techniques that could be applied to better define the relationship between candidate microbes and the development of demyelinating diseases.

## Data Availability

Primary data files are retained by the first and senior authors.

## SUPPLEMENTARY MATERIAL

**Figure S1.**
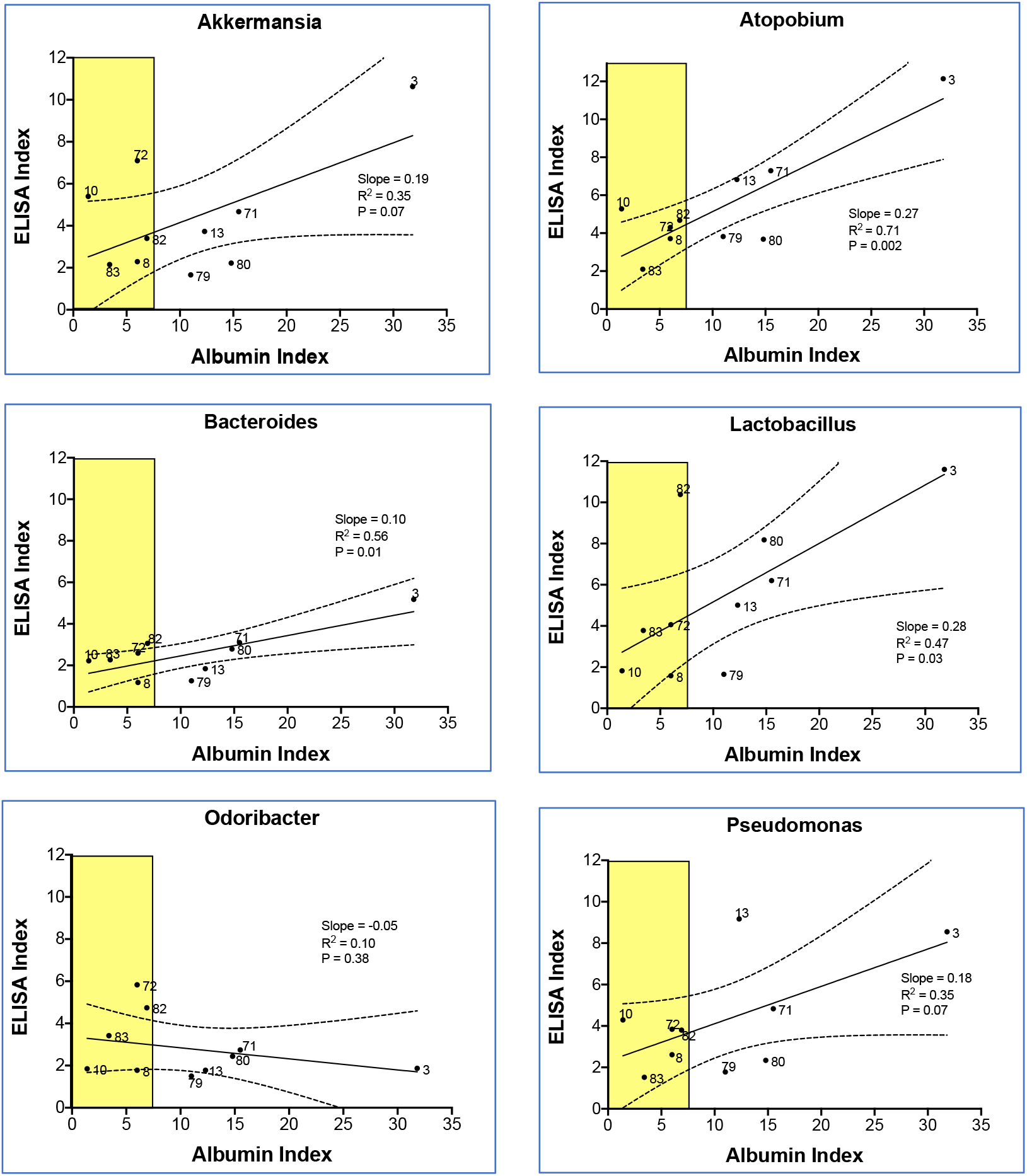
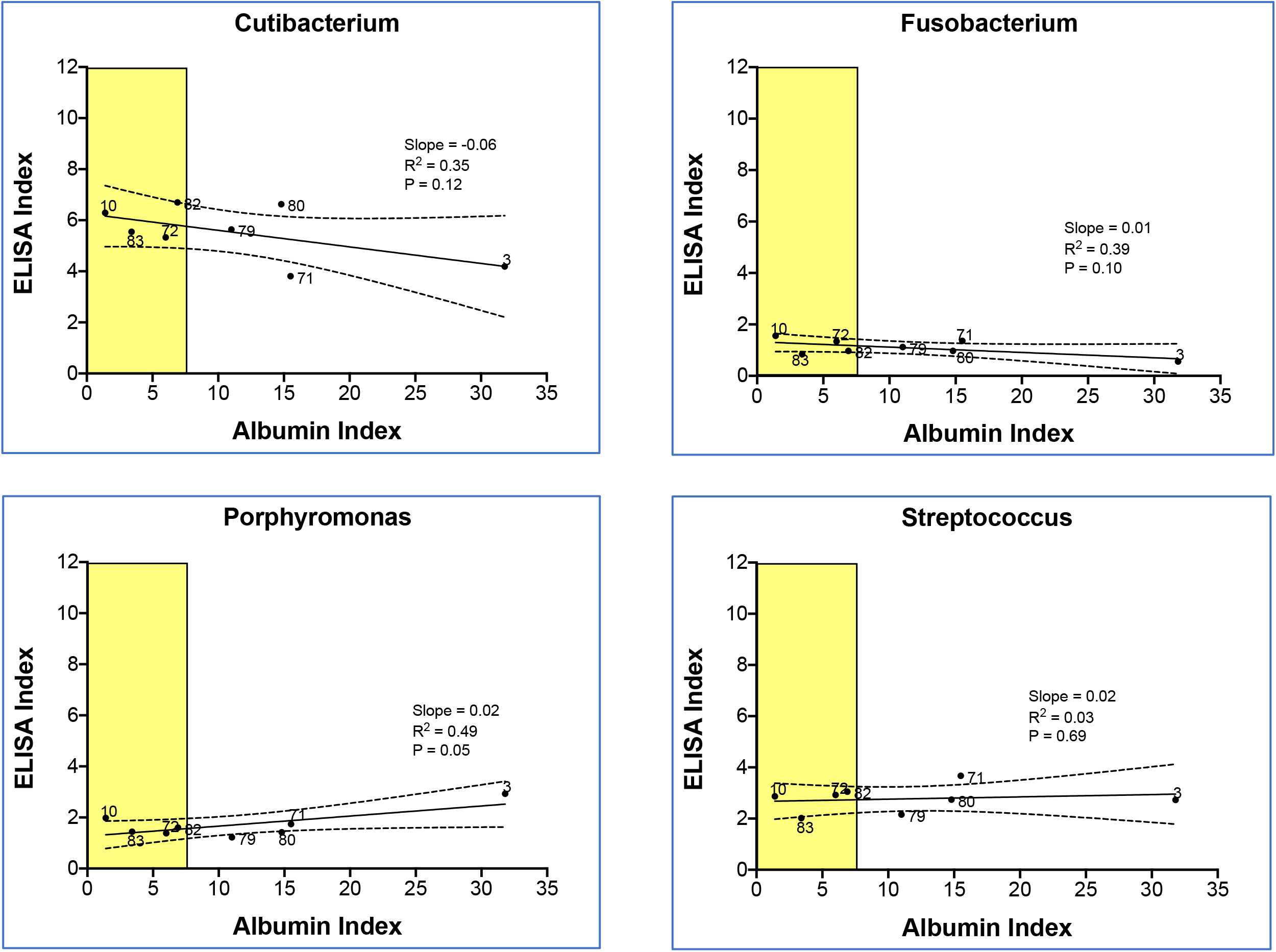
Relationship of ELISA index to blood-brain barrier dysfunction in subjects with demyelinating disease. This exploratory analysis shows the expected ELISA Index (EI) values for a given albumin index (a marker of BBB intactness) for each bacterial antigen. Experimentally determined EI values were plotted against the clinically determined albumin index values, and linear regression was performed (solid lines with dashed 95% confidence bands).^1^ Normal albumin index values (0-9) are indicated by the yellow boxes. Each data point is labeled with its demyelinating disease subject number. More antibodies are expected to leak across the blood-brain barrier (BBB) as the albumin index value rises. Data points above the expected EI for their given albumin index (e.g. subjects 10 and 72 in the anti-Akkermansia plot) are interpreted as evidence for intrathecal antibody synthesis.

**Table S1.** Control and commercial antibodies used for the study.

**A. Control antibody preparations used for indirect ELISA**
| Antibody | Application | Vendor | Catalog number | Clonality | Host | Isotype |
| --- | --- | --- | --- | --- | --- | --- |
| Anti-*E. coli* lipopolysaccharide | primary | Abcam | Ab35654 | Monoclonal | mouse | IgG<sub>2b</sub> |
| Anti-*Pseudomonas aeruginosa* | primary | ThermoFisher | PA1-73116 | Polyclonal | Rabbit | IgG |
| Anti-bacterial Peptidoglycan | primary | EMD Millipore | MAB-995 | Monoclonal | Mouse | IgG<sub>1</sub> |
| HRP anti-mouse IgG | secondary | Vector | PI-2000 | - | Horse | - |
| HRP anti-human IgG | secondary | Jackson Immuno | 309-035-082 | - | Rabbit | - |
| HRP anti-Rabbit IgG | secondary | Vector | PI-1000 | - | Goat | - |

| Organism | Control Primary Ab | Strong + Control Dilution | Weak + Control Dilution | Anti-Rabbit Secondary Ab Dilution | Experimental Human CSF Samples | Secondary Anti-Human Ab Dilution |
| --- | --- | --- | --- | --- | --- | --- |
| Akkermansia | PA1-73116 | 400 | 6,400 | 3,000 | neat | 10,000 |
| Lactobacillus | PA1-73116 | 200 | 1600 | 3,000 | neat | 10,000 |
| Pseudomonas | PA1-73116 | 6,400 | 102,400 | 3,000 | neat | 10,000 |
| Atopobium | PA1-73116 | 400 | 6,400 | 3,000 | neat | 10,000 |
| Bacteroides | PA1-73116 | 200 | 800 | 3,000 | neat | 10,000 |
| Odoribacter | PA1-73116 | 400 | 3,200 | 3,000 | neat | 10,000 |
| Cutibacterium | PA1-73116 | 400 | 3,200 | 6,000 | neat | 20,000 |
| Streptococcus | PA1-73116 | 200 | 400 | 6,000 | neat | 20,000 |
| Porphyromonas | PA1-73116 | 400 | 102,400 | 6,000 | neat | 20,000 |
| Fusobacterium | PA1-73116 | 1,600 | 64,000 | 6,000 | neat | 20,000 |

**Table S2.**
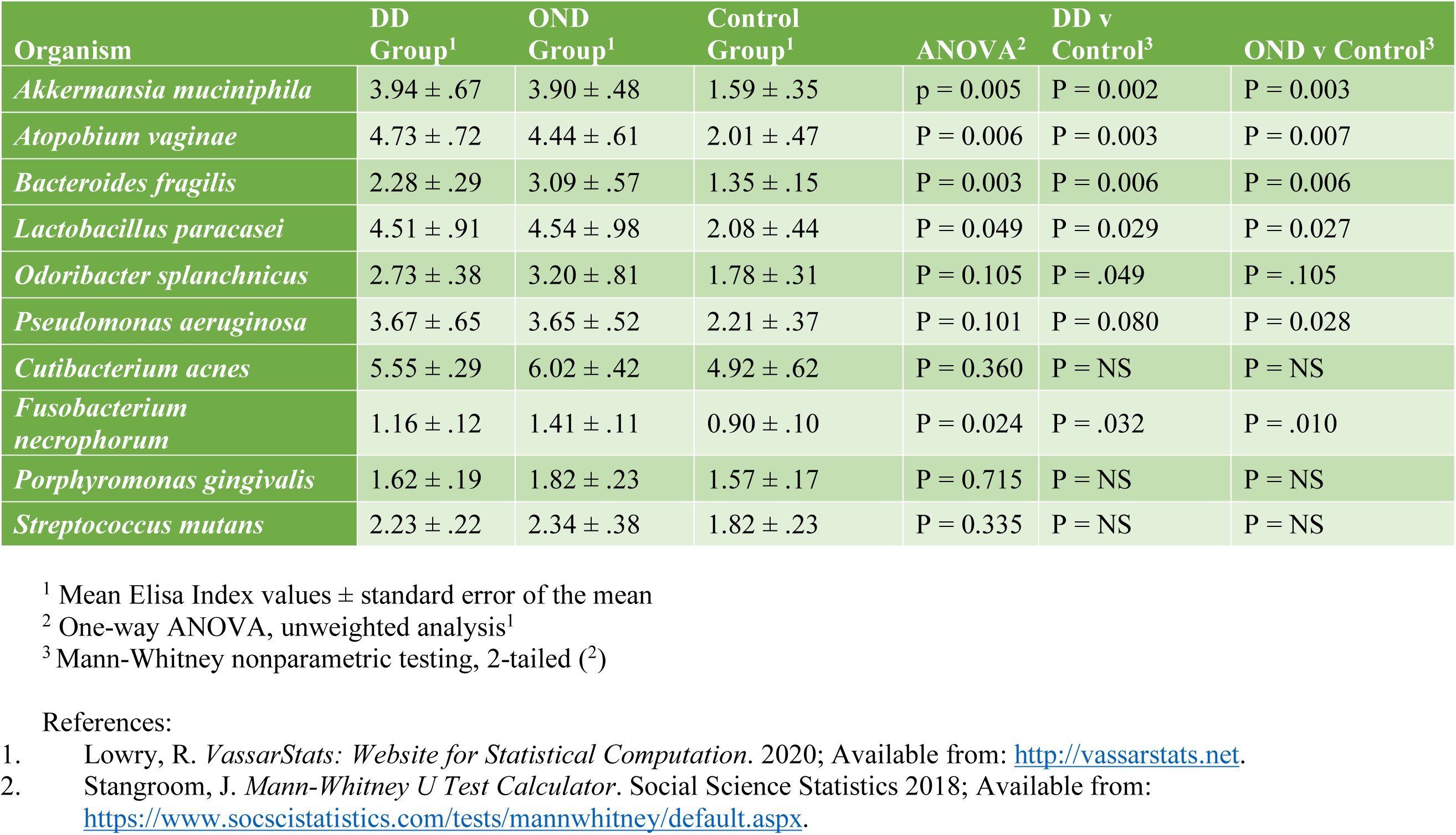
Summary of CSF serologic reactivity and statistical comparisons between the groups.

| Organism | DD Group<sup>1</sup> | OND Group<sup>1</sup> | Control Group<sup>1</sup> | ANOVA<sup>2</sup> | DD v Control<sup>3</sup> | OND v Control<sup>3</sup> |
| --- | --- | --- | --- | --- | --- | --- |
| *Akkermansia muciniphila* | 3.94 ± .67 | 3.90 ± .48 | 1.59 ± .35 | p = 0.005 | P = 0.002 | P = 0.003 |
| *Atopobium vaginae* | 4.73 ± .72 | 4.44 ± .61 | 2.01 ± .47 | P = 0.006 | P = 0.003 | P = 0.007 |
| *Bacteroides fragilis* | 2.28 ± .29 | 3.09 ± .57 | 1.35 ± .15 | P = 0.003 | P = 0.006 | P = 0.006 |
| *Lactobacillus paracasei* | 4.51 ± .91 | 4.54 ± .98 | 2.08 ± .44 | P = 0.049 | P = 0.029 | P = 0.027 |
| *Odoribacter splanchnicus* | 2.73 ± .38 | 3.20 ± .81 | 1.78 ± .31 | P = 0.105 | P = .049 | P = .105 |
| *Pseudomonas aeruginosa* | 3.67 ± .65 | 3.65 ± .52 | 2.21 ± .37 | P = 0.101 | P = 0.080 | P = 0.028 |
| *Cutibacterium acnes* | 5.55 ± .29 | 6.02 ± .42 | 4.92 ± .62 | P = 0.360 | P = NS | P = NS |
| *Fusobacterium necrophorum* | 1.16 ± .12 | 1.41 ± .11 | 0.90 ± .10 | P = 0.024 | P = .032 | P = .010 |
| *Porphyromonas gingivalis* | 1.62 ± .19 | 1.82 ± .23 | 1.57 ± .17 | P = 0.715 | P = NS | P = NS |
| *Streptococcus mutans* | 2.23 ± .22 | 2.34 ± .38 | 1.82 ± .23 | P = 0.335 | P = NS | P = NS |

<sup>1</sup> Mean Elisa Index values ± standard error of the mean
<sup>2</sup> One-way ANOVA, unweighted analysis<sup>1</sup>
<sup>3</sup> Mann-Whitney nonparametric testing, 2-tailed (<sup>2</sup>)

**Table S3.** MS Microbial Candidate List (abridged). Unbiased (deep, next generation) RNA sequencing was performed on 18 formalin-fixed paraffin-embedded primary demyelination brain specimens taken from 17 patients (MS Group), 16 epilepsy brain specimens (Control Group), and 2 blanks (no tissue). This list is revised from that previously published because specimens with gram-negative bacterial sequence (from the sequencing reagents) have been eliminated or resequenced.[1] Methods used for RNA extraction, library preparation, sequencing, and analysis are identical to those previously described, except that MAPQ filtering was not performed here. Criteria for inclusion into this candidate table include:

| UID<sup>1</sup> | Genus | Sum MS Group Reads<sup>2</sup> | Sum Control Group Reads<sup>3</sup> | MS:Control Ratio<sup>4</sup> | Sum Blank Reads<sup>5</sup> | MS:Blank Ratio<sup>6</sup> | # MS Group Specimens where $q < 0.05$<sup>7</sup> |
| --- | --- | --- | --- | --- | --- | --- | --- |
| 239934 | Akkermansia | 25,348 | 314 | 80.7 | 316 | 80.2 | 1 |
| 1380 | Atopobium | 1,426 | 186 | 7.7 | 108 | 13.2 | 1 |
| 816 | Bacteroides | 23,762 | 1,284 | 18.5 | 10,566 | 2.2 | 2 |
| 1578 | Lactobacillus | 166,088 | 23,846 | 7.0 | 9,936 | 16.7 | 1 |
| 283168 | Odoribacter | 2,070 | 28 | 73.9 | 70 | 29.6 | 3 |
| 545932 | Luz24likevirus<sup>8</sup> | 8,338 | 0 | inf | 0 | inf | 5 |
| 1743 | Cutibacterium | 549,878 | 140,666 | 3.9 | 311,544 | 1.8 | 0 |
| 848 | Fusobacterium | 29,446 | 2,320 | 12.7 | 416 | 70.8 | 2 |
| 836 | Porphyromonas | 8,574 | 2,188 | 3.9 | 794 | 10.8 | 0 |
| 1301 | Streptococcus | 570,636 | 44,464 | 12.8 | 54,190 | 10.5 | 4 |

<sup>1</sup> UID = Utah Identification Number (taxon)
<sup>2</sup> Sum of mapped read pairs in the demyelination (MS) group
<sup>3</sup> Sum of mapped read pairs in the control group
<sup>4</sup> (Sum mapped read pairs in the MS group) divided by (Sum mapped read pairs in the Control group)
<sup>5</sup> Sum of mapped read pairs in the blank specimens
<sup>6</sup> (Sum mapped read pairs in the MS group) divided by (Sum mapped read pairs in the blanks)
<sup>7</sup> Number of MS specimens where read-pair mappings to these taxa were significantly increased ( $q < 0.05$ ) compared to the control group
<sup>8</sup> Luz24likevirus is a Pseudomonas phage.
inf = infinity, denominator is zero

- Overexpression in the MS group where
  - the MS:Control Ratio ≥ 3.0
  - the MS:Blank Ratio > 1.5
- > 1000 total reads summed from all 17 specimens in the MS group

